# Twelve-Year Real-World Evaluation of a Regulated Guideline-Based Warfarin Dosing and Care Automation System

**DOI:** 10.64898/2026.08.10.26360059

**Authors:** Mikko T. Tiihonen

## Abstract

**Background:** Warfarin therapy requires repetitive dose adjustments based on INR (International Normalised Ratio) monitoring. We evaluated the long-term real-world performance of Forsante Warfarin Advisor (WA), a CE-marked class IIb guideline-based decision support and care automation medical device used in anticoagulation management.

**Methods:** Retrospective real-world data from routine clinical use between 2014 and 2026 were analysed. Treatment quality was assessed using Time in Therapeutic Range (TTR). Recommendation performance was evaluated by comparing achievement of target INR after clinician acceptance or modification of Warfarin Advisor recommendations.

**Results:** Among 1348 patients in March 2026 median TTR was 83%, compared with 70% in March 2016. Dosages congruent with Warfarin Advisor recommendations were strongly associated with achieving target INR at follow-up in INR target ranges of 2.0-3.0 and 2.5-3.5. Treatment quality remained consistently high across years of deployment. No serious device-attributable safety incidents, regulatory incident reports, or CAPA cases were identified during 12 calendar years and 82,709 patient years of routine use.

**Conclusions:** The findings provide real-world long-term evidence that a guideline-based warfarin dosing and care automation system can support sustained high-quality anticoagulation control in routine clinical practice. They support the feasibility of deploying workflow-integrated execution of selected guideline-driven clinical processes, while the causal effects on clinical outcomes require prospective confirmation.

## 1 Twelve-Year Real-World Evaluation of a Regulated Guideline-Based Warfarin Dosing and Care Automation System

Warfarin remains an important medicine in anticoagulation therapy, especially for patients with mechanical heart valves and other contraindications to direct oral anticoagulants (DOACs). Warfarin has a narrow therapeutic window and serious adverse effects. Frequent laboratory controls of the INR (International Normalised Ratio) are therefore necessary for adjustment of the warfarin dosage. Because a daily dose does not allow for the needed dosing granularity, and the effective biological half-life of warfarin is several days, a weekly schedule is used for dosing. Clinical guidelines, as the one by the Finnish Institute for Health and Welfare (FIHW) (1), have been developed for the warfarin control loop. Traditionally physicians or specially trained nurses have been responsible for dosage adjustment decisions, which were a significant burden before the advent of DOACs in the 2010s.

Warfarin treatment process can be thought of as a control system, where the dosage of warfarin is the control mechanism for the intensity of anticoagulation, and INR tests provide the feedback for the control loop. Due to the long effective half-life of warfarin, there is some inertia to dosage changes. The INR test is usually controlled every three to four weeks in a stable situation but may need to be controlled even daily if the situation is unstable. The target range for INR is 2.0-3.0 in normal warfarin therapy. In the so-called “intensive warfarin therapy” (mostly for patients with mechanical heart valves) the INR target range is 2.5-3.5 (2). Even other target ranges may be used at the clinician’s discretion.

Time in Therapeutic Range (TTR) is commonly used as a quality measure for warfarin therapy. It is calculated from the time that the interpolated INR values have been in the treatment target range, compared to the total time and reported as a percentage (3). TTR values of over 70% have been regarded as indicating high-quality anticoagulation (4).

Computerised systems have been developed for warfarin therapy (5-7). They take care of storing patient data and communicating results to patients. Some also calculate dosage suggestions for the clinician. The published literature focuses mostly on algorithms and short-term evaluation. Evidence for their efficacy in long-term real-world use remains scarce, except for the Poller et al. 2009 study (7).

The Forsante Anticoagulation platform (8) is a comprehensive solution that encodes guideline-derived logic for warfarin dosing. It encompasses the whole process, sending laboratory reminders, dosage instructions, and next control dates to patients using e.g. SMS messages. For stable patients that have their INR values in the INR 2.0-3.0 treatment range, the whole process can be automated within safe limits. The Warfarin Advisor module (9) of the Forsante platform has been CE marked as a class IIb medical device since 2014, and it uses dosing guidance rules derived mainly from the FIHW warfarin dosing guideline (1). Some of the rules were refined using a panel of experts for approval, mainly for resolution of ambiguities. The determination of warfarin dosage is based on the time series of INR values and warfarin dosages.

This study evaluated the long-term real-world performance of Warfarin Advisor as a regulated, guideline-based dosing and care automation system, focusing on population-level treatment quality, recommendation-congruent dosing and subsequent INR control, and safety observations from routine post-market clinical follow-up.

## 2 Methods

### 2.1 Device Description

#### 2.1.1 System and clinical workflow

Warfarin Advisor (WA) is a software module intended for warfarin dosage calculation, CE-marked as a class IIb medical device under applicable EU medical device regulation (UDI-DI: 06429810731006). It encodes the logic in the clinical guidelines by FIHW for warfarin dosing in the stable maintenence phase (1). WA is designed to be incorporated in a software solution to be used by healthcare professionals, and in restricted cases by patients. The intended patient population is adult patients in the stable maintenance phase of warfarin medication, and normal INR target ranges. The WA is to be used by trained clinicians for warfarin dose calculation.

WA is integrated into the Forsante Anticoagulation service, which supports anticoagulation monitoring, treatment guidance, and clinical workflows in a multi-organisation setting. Since the instructions are usually sent to the patient by SMS, the patient has no need for separate mobile apps or a computer, enabling population-wide use of the solution regardless of the patients’ technological proficiency. There is also the possibility of using a responsive mobile web application for input of the results from point-of-care INR testing by the patient or a nurse.

The system supports automated generation and delivery of dosing instructions to eligible patients when INR measurements are within safety limits. Values outside the safe range or situations requiring clinical review are always escalated to a healthcare professional, accompanied by a dosing recommendation, for dosage verification and possible intervention.

#### 2.1.2 Executable guideline logic and validation

The rules in the FIHW guideline have been translated to program logic which uses pattern matching with a decision table, and such program structures that minimise the conceptual distance between the guideline and implementation. Time-weighted linear regression is used for detecting INR trends. A properly balanced and distributed weekly dosage is generated using Shannon entropy and suitable heuristics. Calculation of the Time in Therapeutic Range (TTR) is based on the Rosendaal method, adapted mathematically for computer deployment (3). The implementation has been done in the Racket programming language, currently version 8.5 (10), with total code coverage by tests on unit, integration and system levels.

### 2.2 Evaluated Outcomes

The following outcomes were evaluated:

- Treatment quality as measured by the TTR
- Recommendation performance
- Safety observations
- User reports

### 2.3 Safety surveillance methods

Safety observations concerning WA were gathered from the Forsante ISO 13485 -certified quality management system (QMS) from the 12 years since 2014.

Forsante QMS collects data from customer and internal feedback, communications with authorities, customer follow-up meetings and annual feedback surveys. All feedback is entered into the customer feedback registry or the nonconformity registry, and classified as major or minor, based on possible patient risk. Near misses are classified based on the potential risk. Tickets in the registry are further classified as complaints or other feedback, and complaints are processed further into non-conformity CAPAs, other CAPAs, incidents or change requests in the weekly cross-functional team meetings. Quality manager reviews the feedback and is responsible for the processing.

Forsante Ltd follows the requirements of the EU MDR for the vigilance process. Serious incidents are reported to the competent authority. For non-serious incidents, trends are monitored and reported. The customer feedback and nonconformity registry and possible incidents together with the findings of internal and external audits are reviewed annually, analysed and entered in the structured management review minutes. These minutes were used for safety surveillance analysis, starting from 2014 until the end of 2025, supplemented with the entries in the customer feedback and nonconformity registries after the latest management meeting until the end of March 2026.

The following data was extracted, as pertained to the WA: major customer feedback or nonconformities, CAPAs, incidents with reports to/from regulatory authorities. Additionally, the WA-related software issues with “risk” label were collected from the issue-tracking system.

### 2.4 Data

The study used pseudonymised retrospective data collected under the Forsante QMS. Post-market surveillance and clinical evaluation activities are required under the European Union medical device regulatory framework. The data processing agreement obligations by the healthcare organisations were followed. According to the Finnish requirements and the non-interventional nature of the study, additional ethics committee approval was not required.

### 2.5 Study Design and Analysis Populations

To reduce the possible effect of holiday seasons on the warfarin treatment balance, the month of March was chosen for data extraction. The study comprised three complementary analyses: population-level treatment quality, recommendation-level performance, and post-market safety surveillance:

- **Population-level treatment quality**: repeated cross-sectional TTR snapshots for 2016, 2020, 2024, and 2026.
- **Recommendation-level performance**: March 2026 patients with previous INR within 60 days, analysed by target group.
- **Post-market safety surveillance**: cumulative market exposure of WA over 12 calendar years and 82,709 estimated patient-years.

There were 1348 patients (487 female, 861 male) with an INR measurement during the period of March 2026. The patient selection protocol for primary analyses from that population is in figures 1 and 2.

**Figure 1.**
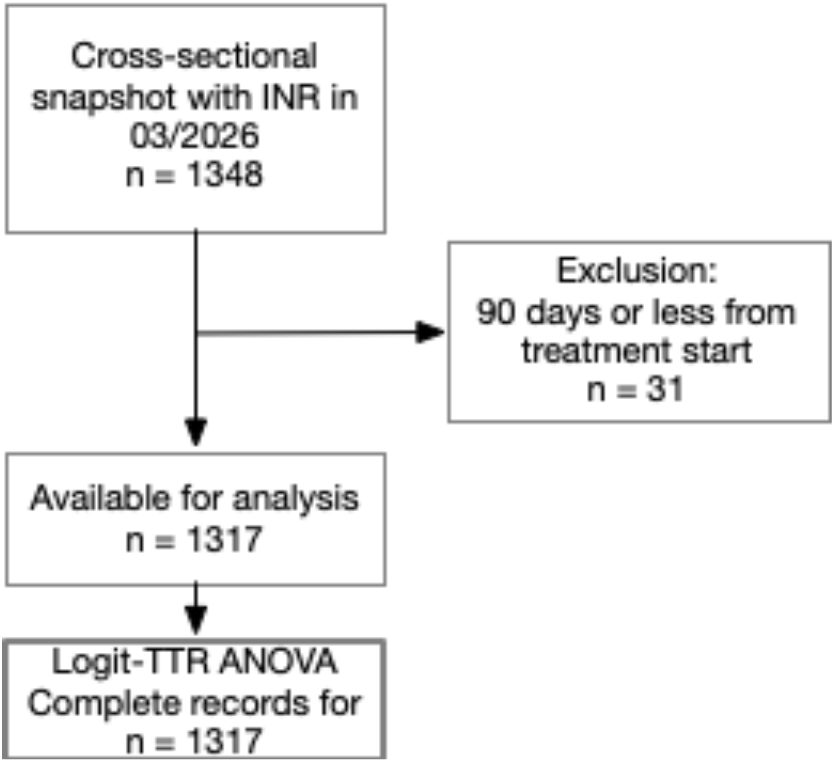
Patient selection for logit-TTR ANOVA

**Figure 2.**
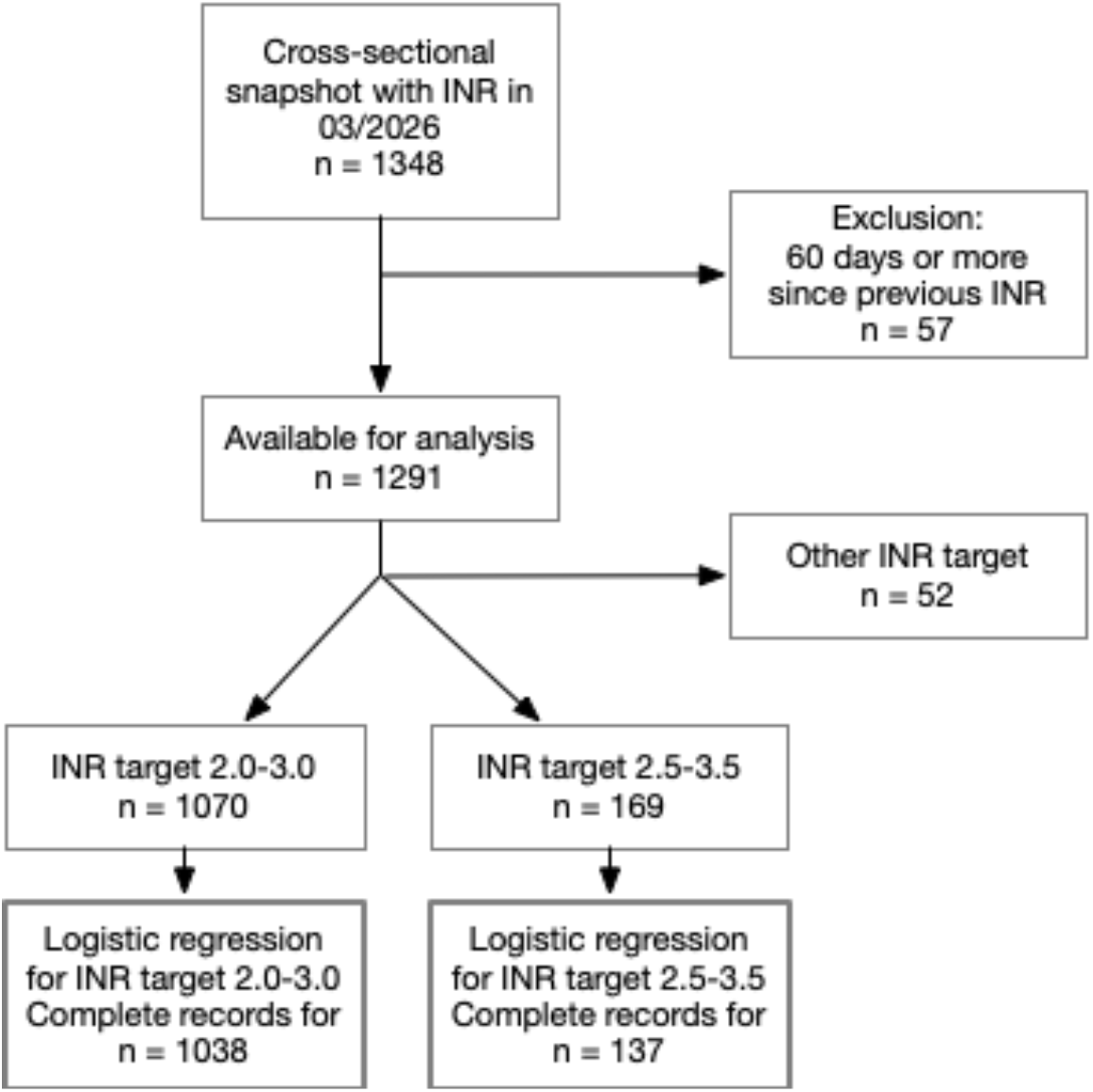
Patient selection for logistic regression for INR-in-target

### 2.6 Statistical Analysis

Descriptive statistics (count, mean, standard deviation [SD], median, interquartile range [IQR]), and contingency tables with Chi-square statistics were used. Ordinary least squares analysis of variance (ANOVA) was used to assess determinants of logit-transformed TTR (with mapping of 0 to 0.001 and 1 to 0.999 before transformation). Logistic regression was used to evaluate achievement of INR in the target range. The recommendation performance of WA was assessed by dividing dosages into two groups: 1. dosages congruent with WA recommendation and 2. dosages deviating from the recommendation.

Predictor selection was guided by collinearity assessment, model diagnostics, and parsimony. Highly correlated variables (r > 0.7) were excluded, potential separation was assessed before model fitting, and continuous predictors were evaluated for linearity in the logit by Lowess-smoothed scatterplots. Non-significant terms were pruned from the model sequentially, handling polynomial terms as entities. Possible confounding factors were retained conservatively and kept if their removal altered the primary effect size by at least 10%. Polynomial predictor terms were retained only when they significantly improved model fit.

Data analysis was conducted using Data Desk, version 8.3 (11).

### 2.7 Reporting

This study is reported in accordance with the Strengthening the Reporting of Observational Studies in Epidemiology (STROBE) guideline (12) (supplementary file).

## 3 Results

### 3.1 Overview of the data

The data set represents routine real-world use of the Forsante Anticoagulation service with WA, encompassing 82,709 patient years (number of dosages in the system multiplied by time in years between dosages, calculated as weighted mean from all cross-sectional samples).

For descriptive statistics of the whole 1348 patient population in March 2026, see Table 1.

**Table 1.**
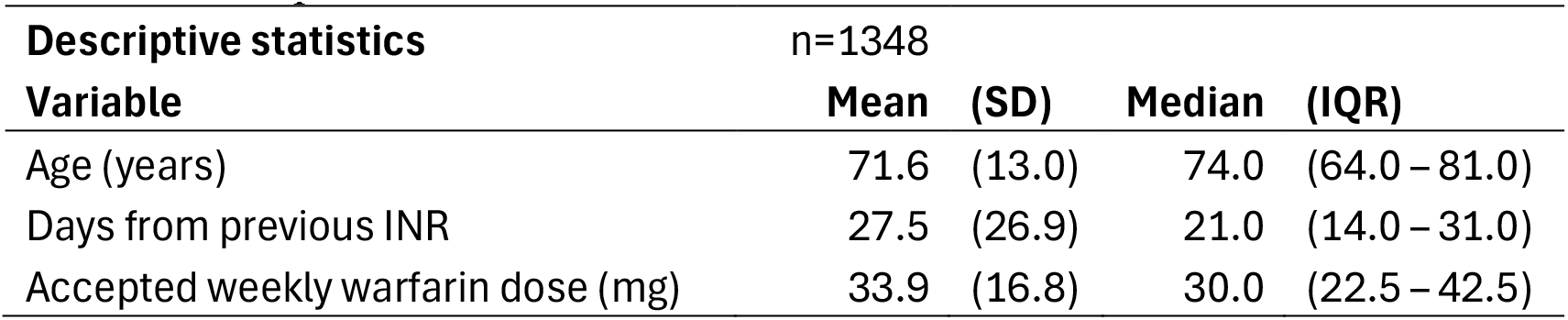
Descriptive statistics for the March 2026 cross section.

| Descriptive statistics |  | n=1348 |  |  |  |
| --- | --- | --- | --- | --- | --- |
| Variable |  | Mean | (SD) | Median | (IQR) |
| Age (years) |  | 71.6 | (13.0) | 74.0 | (64.0 – 81.0) |
| Days from previous INR |  | 27.5 | (26.9) | 21.0 | (14.0 – 31.0) |
| Accepted weekly warfarin dose (mg) |  | 33.9 | (16.8) | 30.0 | (22.5 – 42.5) |

The most common INR-targets used were 2.0-3.0 (83.0%), with a median TTR level of 85% (IQR 74% – 93%), and 2.5-3.5 (13.1%) with median TTR of 75% (IQR 64% – 86%). Other target ranges were infrequent (less than 2% each), lower limit ranging from 1.5 to 3.5 and upper limit from 2.0 to 4.0. Automation was enabled for 812 patients (60.2%).

### 3.2 Treatment quality

Treatment quality was measured by the TTR. To find out how the treatment quality has changed during the lifetime of Forsante Anticoagulation service, the sampling of data was repeated for 2016, 2020, and 2024 in addition to 2026. For this analysis, no exclusions were used. The results are in Table 2.

**Table 2.** Patient age and treatment quality in cross sections.

| Time period | Patients | Mean (SD)<br>age | Mean (SD)<br>TTR | Median<br>TTR | IQR |
| --- | --- | --- | --- | --- | --- |
| March 2016 | 672 | 72.5 (9.7) | 62.4 (34.6) | 70.0% | 31.5% – 94.0% |
| March 2020 | 6472 | 75.0 (11.0) | 79.4 (16.6) | 82.0% | 71.0% – 91.5% |
| March 2024 | 3280 | 73.3 (12.5) | 79.0 (16.9) | 82.0% | 71.0% – 91.0% |
| March 2026 | 1348 | 71.5 (13.0) | 80.1 (16.7) | 83.0% | 72.0% – 92.0% |

An ANOVA model was built for logit-transformed Time in Target Range (logitTTR) using the data for March 2026. Categorical predictors were parameterized using effects coding (sum-to-zero constraints). Candidate predictors were age, sex, INR target range, and automation status. Because automation eligibility depended on INR target range and some combinations were structurally unavailable, INR target range and automation status were combined into a derived categorical variable termed ClinicalProfile. The levels were Normal (INR2.0-3.0), Normal with automation, Intensive (INR 2.5-3.5), Other, and Other with automation. The Pearson correlation matrix did not show any correlations of over 0.7 between the independent predictor variables.

In the logitTTR ANOVA, ClinicalProfile was strongly associated with treatment quality after adjustment for age and sex (F=23.13, p<0.001). Age (F=7.94, p=0.0049) and sex (F=8.23, p=0.0042) were also associated with logitTTR. Parameter estimates are shown in Table 3. The Normal+Automation profile demonstrated the largest positive deviation from the adjusted grand mean (β=1.042, SE=0.188, p<0.0001), followed by the Normal profile (β=0.602, SE=0.199, p=0.0026). The Other profile showed a negative deviation (β=-0.611, SE=0.277, p=0.0276), whereas the Intensive profile was not significantly different from the adjusted grand mean.

**Table 3.** ANOVA for Logit-TTR.

| **Logit TTR** |  |  |  |  |
| --- | --- | --- | --- | --- |
| **Predictor** | **F** | **p** |  |  |
| ClinicalProfile | 23.13 | <0.001 |  |  |
| Sex | 8.23 | 0.004 |  |  |
| Age | 7.94 | 0.005 |  |  |
| **Predictor** | **Coeff.** | **(95% CI)** | **SE** | **p** |
| *ClinicalProfile levels* |  |  |  |  |
| Normal (INR 2.0-3.0) | 0.602 | (0.21 – 0.99) | 0.199 | 0.003 |
| Normal + automation | 1.042 | (0.67 – 1.41) | 0.188 | <0.001 |
| Intensive (INR 2.5-3.5) | -0.2055 | (-0.62 – 0.21) | 0.212 | 0.333 |
| Other | -0.6109 | (-1.15 – -0.07) | 0.277 | 0.028 |
| Other + automation | 0.6562 | (-1.09 – 2.40) | 0.892 | 0.462 |
| *Patient demographics* |  |  |  |  |
| Male sex (vs. female) | 0.309 | (0.10 – 0.52) | 0.108 | 0.004 |
| Age | 0.012 | (0.00 – 0.02) | 0.004 | 0.005 |

*Note: ClinicalProfile was effect-coded (sum-to-zero constraint); coefficients represent deviations from the adjusted grand mean of logitTTR.*

### 3.3 Dosage recommendation performance

The performance of the dosage recommendation can be judged by the INR at the next control measurement: if the INR falls within the target range, the recommendation was considered successful for the purposes of this analysis. This endpoint is an approximation because some patients with substantial initial INR deviations may remain outside target range at the subsequent measurement despite receiving an appropriate dosage adjustment.

In the normal warfarin therapy subgroup (INR 2.0-3.0) 1291 patients were eligible and 1038 were included in the final model for recommendation performance analyses. A logistic regression analysis for the effect of the WA recommendation was performed using the binary response variable “INR-in-target” (INR being in target vs. INR being out of target).

Candidate predictors were acceptance of the WA recommendation (“WA-recommended-dosage”), automation status, age, sex, treatment duration exceeding 90 days (“Treatment-over-90-days”), number of days from previous INR (“Days-from-previous-INR”), previous INR value, and deviation of the previous INR from the midpoint of the target range (“Previous-INR-deviation”). The Pearson correlation matrix showed a high correlation (r>0.7) between previous INR value and Previous-INR-deviation. Continuous variables were checked to have overlap between result categories using boxplots, and categorical variables were checked for non-zero values in contingency tables with INR-in-target (check for “perfect separation”). Previous INR deviation demonstrated a nonlinear association with subsequent INR control, and was therefore represented by linear, quadratic, and cubic terms. Days-from-previous-INR also showed a nonlinear relationship with INR control and was represented by linear and quadratic terms.

The final model (Table 4, Fig. 3, Fig 4.) retained WA-recommended-dosage, treatment-over-90-days, sex, previous-INR-deviation terms, and days-from-previous-INR terms.

**Table 4.** Logistic regression analysis for INR hitting the 2.0-3.0 target.

| INR in target |  | Target 2.0-3.0 |  |  |  |
| --- | --- | --- | --- | --- | --- |
| Predictor | Odds ratio | (95% CI) | Coeff. | SE | p |
| WA-recommended-dosage | 35.0 | (21.9 – 55.8) | 3.554 | 0.169 | <0.001 |
| Treatment-over-90-days | 3.3 | (1.8 – 6.0) | 1.197 | 0.217 | 0.006 |
| Male sex | 1.5 | (1.2 – 1.9) | 0.420 | 0.086 | 0.014 |
| Previous-INR-deviation | *Nonlinear term* |  | 0.321 | 0.210 | 0.126 |
| Previous-INR-deviation<sup>2</sup> | *Nonlinear term* |  | -1.127 | 0.327 | 0.001 |
| Previous-INR-deviation<sup>3</sup> | *Nonlinear term* |  | 0.337 | 0.141 | 0.017 |
| Days-from-previous-INR | *Nonlinear term* |  | 0.056 | 0.029 | 0.054 |
| Days-from-previous-INR<sup>2</sup> | *Nonlinear term* |  | -0.001 | 0.001 | 0.049 |

**Figure 3.**
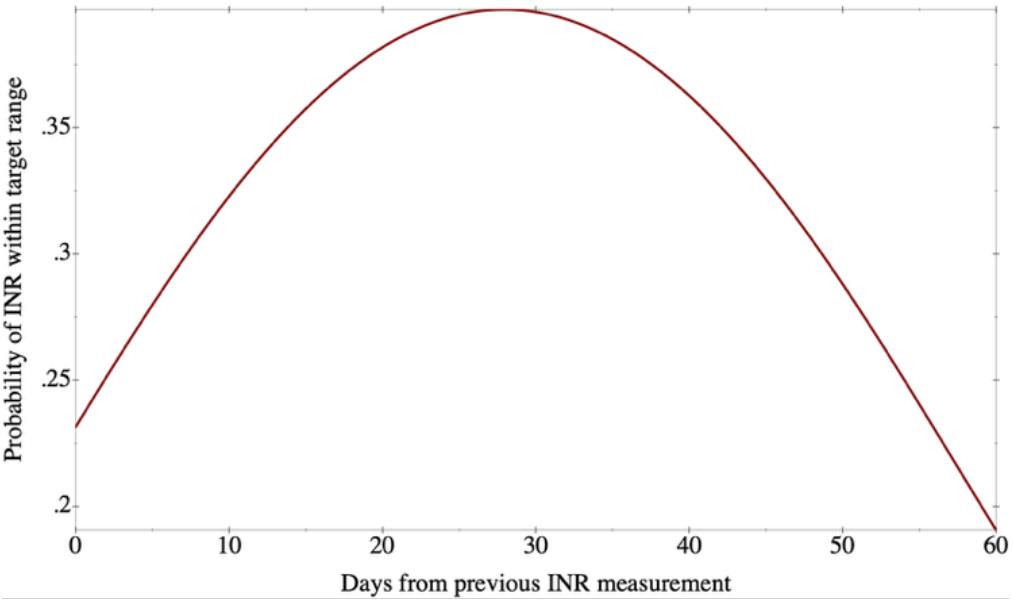
The polynomial effect of Days-from-previous-INR on probability of INR-in-target (2.0-3.0)

**Figure 4.**
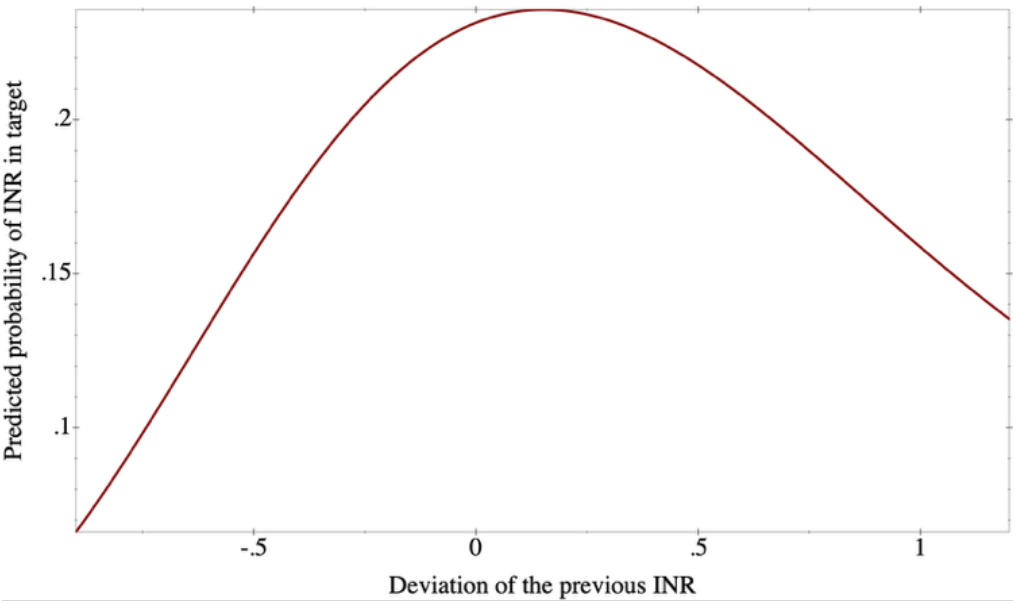
The polynomial effect of Previous-INR-deviation on probability of INR-in-target (2.0-3.0)

WA-recommended-dosage was the strongest predictor of achieving target INR (OR 35.0, 95% CI 21.9–55.8, p<0.001). Treatment-over-90-days (OR 3.3, 95% CI 1.8–6.0, p=0.006) and male sex (OR 1.5, 95% CI 1.2–1.9, p=0.014) were also independently associated with successful INR control.

In the intensive warfarin therapy subgroup (INR target 2.5-3.5) 169 patients were eligible and 137 were included in the final model for recommendation performance analyses. The Pearson correlation matrix showed a high correlation (r>0.7) between previous INR and Previous-INR-deviation. Continuous variables were checked to have overlap between result categories using boxplots, and categorical variables were checked for non-zero values in contingency tables, revealing zero cells for treatment-over-90-days vs. INR-in-target. Previous INR deviation demonstrated a nonlinear association with subsequent INR control, and was represented by linear, quadratic, and cubic terms. Days-from-previous-INR also showed a nonlinear relationship with INR control and was represented by linear and quadratic terms. WA-recommended-dosage and Previous-INR-deviation were retained in the model.

Results (Table 5, Fig. 5) were similar as for the normal warfarin therapy subgroup; the WA-recommended-dosage showing a marked effect (OR 11.3, CI 5.5-23.4, p<0.001), with substantially higher odds of INR control in the group with WA recommendation acceptance as compared with non-acceptance. The cubic polynomial of Previous-INR-deviation was also significantly associated with INR control. In this smaller group, the other factors did not reach statistical significance and were thus not included in the model.

**Table 5.**
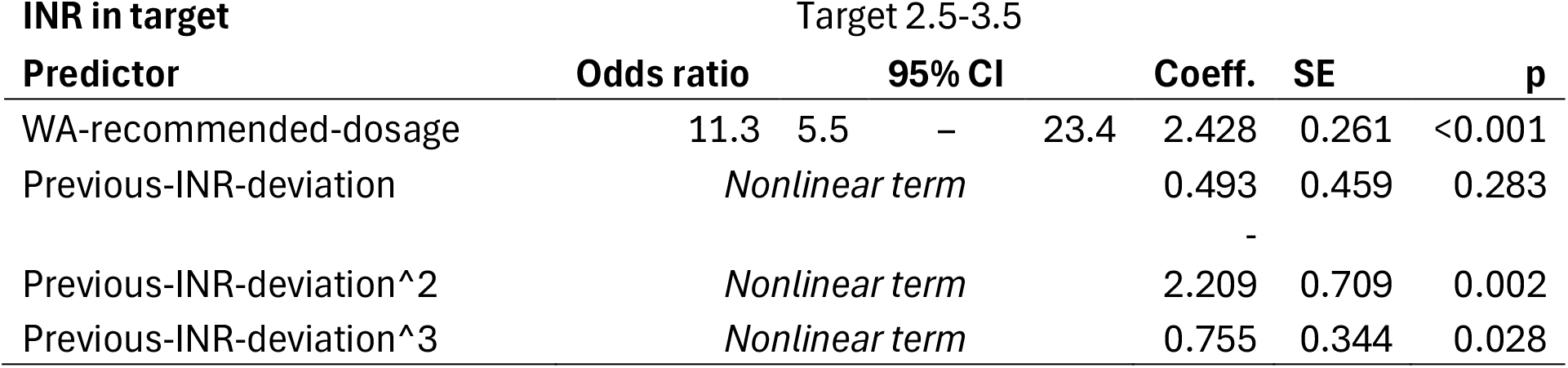
Logistic regression analysis for INR hitting the 2.5-3.5 target.

**Figure 5.**
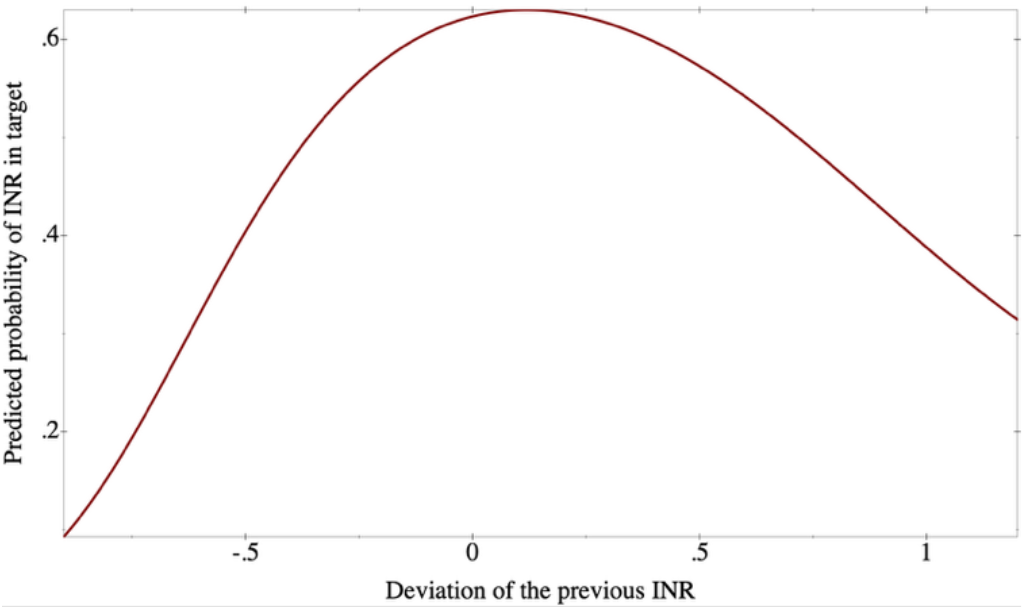
The polynomial effect of Previous-INR-deviation on probability of INR-in-target (2.5-3.5)

In a sensitivity analysis using absolute INR deviation as the outcome, WA recommendation congruence, absolute previous INR deviation and time from previous INR were statistically significant predictors. Full model results are provided in the Appendix A.

As automation bias might affect the choice of dosage by the professional, we performed an explanatory analysis to examine the success of dosages given without any WA recommendation. This situation may arise when the INR is not received from the laboratory information system in due time for technical reasons, and the clinician enters the INR (from the EHR) and dosage manually. In the dataset from 2020 there were the largest number of such cases after the initial period, 110 (out of total 6472 cases). In that dataset, the contingency table of INR being in target, explained by the availability of recommendation, showed that the proportion of successful dosages (“hit rate”) was 62.7% without the recommendation and 81.2% with the WA recommendation (Chi-square 23.7, 1 df, p<0.001).

### 3.4 Safety results

Safety results are summarised in Table 6. No major customer feedback was received for WA. No CAPA issues concerning the WA have been filed during its time on market and no patient adverse effects have been reported. No major incidents concerning WA were filed or reported to regulatory authorities. Seven “risk” labelled software issues have been detected and resolved. Of these seven, three originated in customer complaints (related to the distribution of the weekly dose), four were of internal origin.

**Table 6.**
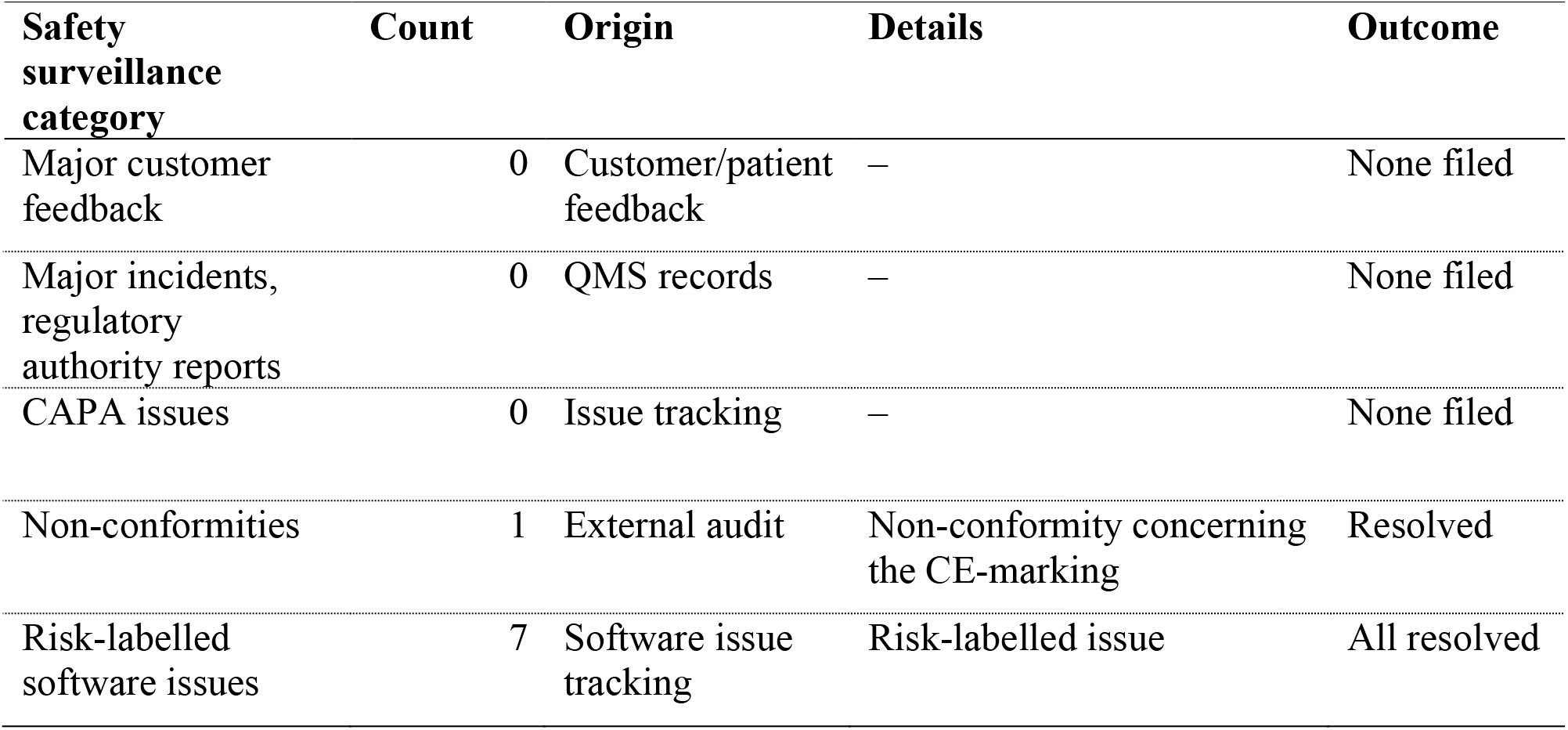
WA safety surveillance data from 2014-2026.

| **Safety surveillance category** | **Count** | **Origin** | **Details** | **Outcome** |
| --- | --- | --- | --- | --- |
| Major customer feedback | 0 | Customer/patient feedback | – | None filed |
| Major incidents, regulatory authority reports | 0 | QMS records | – | None filed |
| CAPA issues | 0 | Issue tracking | – | None filed |
| Non-conformities | 1 | External audit | Non-conformity concerning the CE-marking | Resolved |
| Risk-labelled software issues | 7 | Software issue tracking | Risk-labelled issue | All resolved |

## 4 Discussion

### 4.1 Principal findings

Forsante Anticoagulation service with WA was associated with sustained treatment quality and there were no identified serious device-attributable safety concerns during post-market surveillance. The treatment quality observed in the Forsante Anticoagulation population was high. The observed rise in median TTR from 70.0% in March 2016 to 83.0% in March 2026 indicates improved anticoagulation quality over the evaluated deployment period. The median TTR values in both normal and intensive warfarin therapy subgroups (85% and 75%, respectively) were on a good level. The clinical profile with normal INR target range and automation was associated with the largest positive deviation towards higher TTR values.

Dosages congruent with WA recommendations were associated with substantially higher odds of subsequent INR-in-target status both in normal and intensive warfarin therapy. There was further support for this, since the dosages provided by the professional without WA recommendation achieved the target only in 62.7% of cases, while the success rate for WA congruent dosages was 81.2%.

These findings support the real-world performance of a guideline-based dosing algorithm sustaining high-quality warfarin management. Because recommendation acceptance was not randomised and clinician overrides may have occurred in more complex cases, these findings should be interpreted as a real-world performance signal rather than as a causal estimate of algorithmic efficacy.

For patients that need to use warfarin because of contraindications to DOACs, such as mechanical heart valves, being able to achieve a high TTR is an important benefit.

### 4.2 Comparison with prior literature

As compared to WA, other solutions’ mean TTR values in the literature have been lower, from 59% to 74% (5, 13, 14).

The significance of using recommendation-congruent dosages is in accord with the previous findings showing improved performance of algorithmic dosing over manual dosing (15, 16). The WA 81.2% “hit rate” in achieving INR values in the target range is better than that of the AuriculA system (72%), which was deployed in a similar socio-economic setting (15).

### 4.3 Implications for clinical informatics

As WA encodes guideline-derived logic for care execution, these findings may have implications beyond anticoagulation management. Clinical guidelines have the potential to reduce clinical variation and thus improve healthcare quality (17). The bottleneck has been in the execution. The estimated workload for comprehensive execution of clinical guidelines in a general practice exceeds human capabilities (18).

WA demonstrates how structured guideline-derived clinical knowledge can be translated into executable decision logic within routine healthcare workflows. The successful long-term operation of a guideline-based dosing system in a high-stakes clinical setting provides evidence supporting further evaluation of similar approaches in other guideline-driven clinical domains. In such systems, the principal challenge is no longer the availability of clinical knowledge, but its reliable and scalable execution within routine care pathways.

Unlike traditional clinical decision support systems that primarily provide recommendations for clinician interpretation, WA operationalises bounded guideline-based decisions under predefined governance and escalation rules. Distinguishing between decision support and decision execution approaches may therefore provide a useful conceptual framework for discussing clinical automation.

Importantly, the present study does not suggest that computerised systems widely replace clinical judgement. Rather, it supports the idea that selected repetitive clinical processes may be automated using guideline-concordant software-generated decisions. Clinical expertise will always be needed in exceptional situations. It is the responsibility of the software manufacturer to ensure that such situations are identified and escalated to the clinician. Determining whether a patient is suitable for such guideline-aligned automation remains a clinical responsibility which cannot be delegated. In many cases that determination can be made, opening the possibility for significant improvements in treatment efficiency and quality.

### 4.4 Limitations

The current study is retrospective and non-randomised; therefore, all the usual limitations of observational studies need to be considered (19, 20).

The clinically most significant endpoints, major bleeding, thromboembolic complications and death were not accessible to the system and thus could not be analysed. TTR is a surrogate measure for these risks (21).

Since the usage of warfarin in Finland has declined steadily over the past ten years, as patients have been changed to DOACs, the possibility of a selection or survivor bias needs to be considered. This bias could have contributed to the observed increase in population-level TTR over time, as patients with poor adherence might have been switched to DOACs, leaving the patients with better treatment balance and higher TTR values. However, this would not have affected the success rate of WA generated vs. manual dosages. The mean age of patients remained quite similar at the evaluated time points, which does not indicate bias.

It is possible, that clinicians could have overridden the WA recommendations in difficult clinical situations, thereby affecting the success of manual dosing. Because no randomised comparator group existed, causal conclusions regarding the effect of WA cannot be drawn. Automation eligibility was determined by clinical criteria rather than random allocation. Because cross-sectional analyses were based on March data, the results may not capture seasonal variation or changes in treatment quality during other parts of the year.

Safety observations were based on events captured in the quality management system and therefore may not include unreported clinical events or events not attributed to the system by users or healthcare organisations. The study comprised only patients from Finnish healthcare organisations, which affects the generalisability to countries with lower standards of living and healthcare.

### 4.5 Conclusions

In this long-term real-world evaluation, use of a regulated guideline-based warfarin dosing and care automation system was associated with sustained high TTR levels in routine anticoagulation care. Dosages aligned with Warfarin Advisor recommendations were associated with higher rates of subsequent INR control than clinician-modified dosages, both in normal and intensive warfarin therapy. Across 12 calendar years and 82,709 estimated patient-years of use, no serious device-attributable safety incidents, regulatory incident reports, or CAPA cases concerning Warfarin Advisor were identified in the quality management system.

Because the study was retrospective and non-randomised, causal conclusions on algorithmic efficacy and clinical outcome effects cannot be ascertained. Nevertheless, the findings support the feasibility of bounded, workflow-integrated guideline execution for selected repetitive clinical processes, with exceptional cases escalated to healthcare professionals.

Future research should investigate the applicability of guideline-based clinical automation approaches in other therapeutic domains and evaluate their long-term clinical and economic impact in a randomised setting.

## Supporting information

STROBE checklist

## Data Availability

The underlying patient-level data contain protected health information and cannot be publicly shared. Aggregate data supporting the findings may be available from the corresponding author upon reasonable request and subject to applicable legal and contractual restrictions.

## Declaration of Generative AI and AI-Assisted Technologies in the Writing Process

During the preparation of this manuscript, the author used Microsoft Copilot as a writing assistance tool for proofreading, language editing, and feedback on manuscript structure, clarity, and organization. The tool was not used to generate, analyse, interpret, or modify research data, nor was it used for statistical analyses or scientific conclusions. All suggestions provided by the tool were critically reviewed by the author and adopted only when considered scientifically appropriate. The final manuscript was independently written, verified, and approved by the author, who assumes full responsibility for the accuracy, integrity, and content of the work.

## Acknowledgements

BSc Kasper Kautto helped in data pseudonymisation and retrieval, which is gratefully acknowledged.

## Appendix A

### INR deviation change analysis of variance

#### Population

All 1291 patients available for analysis (see fig. 2), encompassing all INR targets.

#### ANOVA

INR deviation change at the time of March 2026 was calculated as the absolute value of the subtraction of two distances from the target range midpoint: The deviation of the previous INR (“prev-INR-deviation”) was subtracted from deviation of the index INR value of March 2026.

An analysis of variance model using ordinary least squares (OLS) was built for INR-deviation using the data for March 2026. Candidate predictors were acceptance of the WA recommendation (“WA-recommended-dosage”), prev-INR-deviation, age, sex, treatment duration exceeding 90 days (“treatment-over-90-days”), number of days from previous INR (“Days-from-previous-INR”) and target INR range. Because automation eligibility depended on INR target range and some combinations were structurally unavailable, INR target range and automation status were combined into a derived categorical variable termed ClinicalProfile. The levels were Normal (INR2.0-3.0), Normal with automation, Intensive (INR 2.5-3.5), Other, and Other with automation. The Pearson correlation matrix did not show any correlations of over 0.7 between the independent predictor variables. Categorical predictors were parameterized using effects coding (sum-to-zero constraints). Normal probability plots of continuous predictors were visually checked for normality violations, and prev-INR-deviation was transformed logarithmically. The Pearson correlation matrix did not show any correlations of over 0.7 between the independent predictor variables.

The final model retained WA-recommended-dosage, logarithm (base 10) of prev-INR-deviation, days-from-previous-INR, and treatment-over-90-days (Table A1).

**Table A1.** ANOVA for INR deviation change.

| INR deviation change |  |  |  |  |
| --- | --- | --- | --- | --- |
| Predictor | Coeff | (95% CI) | SE | p |
| WA-recommended-dosage used | -0.327 | (-0.409 – -0.266) | 0.037 | <0.001 |
| log(prev-INR-deviation) | -0.557 | (-0.614 – -0.498) | 0.030 | <0.001 |
| days-from-previous-INR | 0.002 | (0.001 – 0.005) | 0.001 | 0.013 |
| treatment-over-90-days | -0.002 | (-0.001 – 0.144) | 0.001 | 0.068 |

